# The impact of 22q11.2 copy number variants on human traits in the general population

**DOI:** 10.1101/2022.09.21.22280207

**Authors:** Malú Zamariolli, Chiara Auwerx, Marie C Sadler, Adriaan van der Graaf, Kaido Lepik, Mariana Moysés-Oliveira, Anelisa G Dantas, Maria Isabel Melaragno, Zoltán Kutalik

**Affiliations:** Genetics Division, Universidade Federal de São Paulo, São Paulo, Brazil; Department of Computational Biology, University of Lausanne, Lausanne, Switzerland; Swiss Institute of Bioinformatics, Lausanne, Switzerland; University Center for Primary Care and Public Health, University of Lausanne, Lausanne, Switzerland; Center for Integrative Genomics, University of Lausanne, Lausanne, Switzerland

**Author notes:** **Corresponding author:** Zoltán Kutalik, Department of Computational Biology, University of Lausanne, Lausanne, Switzerland., Swiss Institute of Bioinformatics, Lausanne, Switzerland., University Center for Primary Care and Public Health, University of Lausanne, Lausanne, Switzerland.

## Abstract

While extensively studied in clinical cohorts, the phenotypic consequences of 22q11.2 copy number variants (CNVs) in the general population remain understudied. To address this gap, we performed a phenome-wide association scan in 405’324 unrelated UK Biobank (UKBB) participants using CNV calls from genotyping array. We mapped 236 Human Phenotype Ontology terms linked to any of the 90 genes encompassed by the region to 170 UKBB traits and assessed the association between these traits and the copy-number state of 504 SNP-array probes in the region. We found significant associations for eight continuous and nine binary traits associated under different models (duplication-only, deletion-only, U-shape and mirror model). The causal effect of the expression level of 22q11.2 genes on associated traits was assessed through transcriptome-wide mendelian randomization (TWMR), revealing that increased expression of *ARVCF* increased BMI. Similarly, increased *DGCR6* expression causally reduced mean platelet volume, in line with the corresponding CNV effect. Furthermore, cross-trait multivariable mendelian randomization (MVMR) suggested a predominant role of genuine (horizontal) pleiotropy in the CNV region. Our findings show that within the general population, 22q11.2 CNVs are associated with traits previously linked to genes in the region, with duplications and deletions acting upon traits in different fashion. We also showed that gain or loss of distinct segments within 22q11.2 may impact a trait under different association models. Our results have provided new insights to help further the understanding of the complex 22q11.2 region.

## INTRODUCTION

The 22q11.2 region is a structurally complex region of the genome due to the presence of segmental duplications or low copy repeats (LCRs), named LCRA to LCRH, which predispose the region to genomic rearrangements, resulting in deletions or duplications of different segments. Specifically, deletions within the ∼3 Mb segment from LCRA to LCRD represent the main cause of the 22q11.2 deletion syndrome (22q11.2DS), the most frequent microdeletion syndrome in humans, with an estimated incidence between 1: 3000 and 1: 6000 live births ^1^.

Studies in clinical cohorts have investigated the phenotypic consequences of the 22q11.2 deletion, which include cardiac defects, facial and palate alterations, immunodeficiencies, endocrine, genitourinary, and gastrointestinal alterations ^1,2^, developmental delay, cognitive deficits, and psychiatric disorders, such as schizophrenia ^1^. In contrast, the phenotypic consequences of the region’s duplication remain more elusive. Most of what is known is based on studies of a few individuals or families, but the findings indicate pleiotropy and variable consequences, similarly to the deletion. Some features, such as heart defects, velopharyngeal insufficiency, and neurodevelopmental and psychiatric disorders are shared with the 22q11.2DS ^3,4^. Other 22q11.2 duplication carriers exhibit very mild or unnoticeable phenotypes ^5^, suggesting variable expressivity and/or reduced penetrance. Finally, rare single nucleotide variants (SNVs) in genes encompassed by the region have been linked to various disorders, such as Bernard–Soulier syndrome, caused by SNVs in *GP1BB* ^6^, or CEDNIK syndrome, caused by SNVs in *SNAP29* ^7^. Overall, the multitude of variants and phenotypes that have been linked to the 22q11.2 LCRA to LCRD region highlights its clinical relevance. Because of their highly deleterious impact, 22q11.2 variants are often investigated in clinical settings. Studied cohorts are thus heavily biased towards individuals with severe phenotypic manifestation, leading to an incomplete and biased understanding of these variants’ role in the human population. This is particularly relevant considering recent studies that have shown variable expressivity and incomplete penetrance of SNVs ^8,9^ and CNVs ^10^ that were previously believed to be highly pathogenic, including at the 22q11.2 LCRA-LCRD locus ^11^. To address this gap, we performed a phenome-wide analysis in the UK Biobank (UKBB) (Bycroft et al., 2018), a populational cohort of ∼500,000 individuals, to identify associations of 22q11.2 CNVs with traits previously implicated by their genetic content.

## MATERIAL AND METHODS

### Cohort description

Analyses were performed in the UK Biobank (UKBB), a volunteer-based cohort from the general UK adult population (Bycroft et al., 2018). Gender mismatched, related and retracted samples (by 09/08/2021), as well as CNV outliers (see **CNV calling**) were excluded, resulting in a total of 405’324 participants (218’719 females and 186’605 males) used for the analyses. Individuals were aged between 40 and 69 years at recruitment. All participants signed a broad informed consent form and data was accessed through a UKBB application (#16389).

### 22q11.2 Region Definition

The 22q11.2 region was defined as chr22:18,630,000_21,910,000 based on the human genome reference build GRCh37/hg19 in order to encompass LCRs from A to D. The 90 NCBI RefSeq genes contained in the region were downloaded from the UCSC Table Browser (http://genome.ucsc.edu/cgi-bin/hgTables?command=start).

### Trait Selection

Phenotypes linked to the 22q11.2 region’s genetic content were identified using the Human Phenotype Ontology (HPO) mapping ^13^, an ontology-based system that uses information from different medical sources including OMIM and Orphanet. Genes and their most specific associated HPO term (i.e., not all ancestors) were downloaded from the HPO database (http://purl.obolibrary.org/obo/hp/hpoa/genes_to_phenotype.txt - Accessed on 22/10/2021). Overall, 24 out of 90 genes in the 22q11.2 region were associated to at least one HPO term, yielding 631 associated HPO terms.

### Mapping of HPO terms to UKBB binary traits

To map HPO terms to binary UKBB traits, two complementary approaches were used. First, the online tool EMBL-EBI Ontology Xref Service (OxO) (https://www.ebi.ac.uk/spot/oxo/) was used to map HPO terms to International Classification of Diseases, 10^th^ Revision [ICD-10] codes, followed by manual curation and grouping of ICD10 codes into broader phenotypes when appropriate according to the Phecode map ^14^. Remaining HPO terms were mapped to Phecode definitions using manual curation by Bastarache et al. (2018)^15^. Mapping was manually curated and only phenotypes with at least 500 cases were retained. In addition, individuals with a related ICD10 code or self-reported disease to the one studied were excluded from controls in a phenotype-specific fashion (**Table S1**). Overall, 218 HPO terms were mapped to 152 UKBB binary traits (**Table S2**). The number of individuals by phenotype is reported in **Table S3**.

### Mapping of HPO terms to UKBB continuous traits

An in-house web scraping approach was developed to map HPO terms to UKBB continuous traits. A list of 1’769 continuous UKBB measures was used as input on the HPO database (https://hpo.jax.org/app/) to obtain the web page’s results for each query. Results were filtered for HPO terms of interest i.e., 631 terms linked to 22q11.2 genes. With this approach, 18 UKBB continuous traits were obtained from 18 HPO terms (**Table S4**). The number of individuals by trait is reported in **Table S5**.

### 22q11.2 CNV association scan

#### CNV calling

CNVs were called with PennCNV v1.0.5 and underwent quality control as described in Auwerx et al., 2022. Briefly, a quality score reflecting the probability for the CNV to be a true positive was assigned to each call and used for filtering (|QS| ≥ 0.5) ^16^. CNVs from samples genotyped on plates with a mean CNV count per sample > 100 or from samples with > 200 CNVs or a single CNV > 10 Mb were excluded to minimize batch effects, genotyping errors, or extreme chromosomal abnormalities.

CNV calls were transformed into probe-by-sample matrices with copy-number state for each probe (deletion = -1; copy-neutral = 0; duplication = 1).

#### Plink encoding and association models

Probe-level matrices were converted to PLINK binary file sets, with copy-number states being encoded to accommodate analysis according to four different association models: duplication-only, deletion-only, mirror, and U-shape model (**Table 1**). The duplication-only model assessed the impact of duplications disregarding deletions; the deletion-only model assessed the impact of deletions disregarding duplications; the mirror model assessed the additive effect of each additional copy of a probe (i.e., duplications and deletions have opposing effects); while the U-shape model assumes that duplications and deletions have the same effect direction ^10^.

**Table 1.** PLINK encoding of CNVs into association models.

| **Association Model** | **Deletion-only** | **Duplication-only** | **Mirror** | **U-shape<sup>a</sup>** |
| --- | --- | --- | --- | --- |
| **Deletion** | TT | 00 | AA | AA |
| **Copy-Neutral** | AT | AT | AT | AT |
| **Duplication** | 00 | TT | TT | TT |

<sup>a</sup>For the U-shape model, the “hetonly” modifier in Plink was used.

#### CNV probe selection and number of effective tests

Probes with high genotype missingness (> 5%) were excluded, resulting in 864 CNV proxy probes spanning chr22:18,630,000_21,910,000. We retained 504 CNV proxy probes that are highly correlated (r^2^ ≥ 0.999) to at least ten other probes, allowing to reduce the multiple testing burden while ensuring that selected probes adequately capture the CNV landscape of the region.

The number of effective probes (i.e., number of probes required to capture 99.5% of the variance in the probe-by-sample CNV matrices) was calculated according to Gao et al. (2008) based on the 504 CNV proxy probes (N_eff-probes_ = 6). The same approach was used to account for correlation among 18 continuous (N_eff-continuous_ = 16) and 152 binary traits (N_eff-binary_ = 113). This resulted in 774 effective tests (N_eff_ = N_eff-probes_ * (N_eff-continuous_ + N_eff-binary_)), setting the threshold for significance at p ≤ 0.05/774 = 6.5 × 10^−5^.

#### Continuous traits

The 18 selected continuous traits were inverse normal transformed and corrected for covariates: age, age^2^, sex, genotyping batch, and principal components (PCs) 1–40. Associations between the copy number (CN) of selected probes and normalized covariate-corrected traits were performed in PLINK v2.0 according to all four association models using linear regression, as previously described ^10^. Significant associations (p ≤ 6.5 × 10^−5^) were retained.

#### Binary traits

For each trait, covariates among age, age^2^, sex, genotyping batch, and principal components (PCs) 1–40 that were significantly associated with the trait (p ≤ 0.05) were selected with logistic regression in R. Associations between the copy number (CN) of selected probes and 152 binary selected traits were performed in PLINK v2.0 according to all four association models using logistic regression and correcting for trait-specific selected covariates. Significant associations (p ≤ 6.5 × 10^−5^) were retained.

#### Stepwise conditional analysis

The number of independent signals per trait and association model was determined by stepwise conditional analysis ^10^, i.e., CNV status of the lead probe was regressed out from the trait and association scan was conducted again until no more significantly associated probes remained.

#### Sensitivity analysis

Due to the low frequency of CNVs within the 22q11.2 region, alternative tests were performed to ensure the confidence of significant associations. For significant associations with continuous traits, a Wilcoxon rank-sum test was performed as a sensitivity analysis to assess agreement with linear regression. Significant associations with binary traits were retained only when confirmed by at least one of two approaches: 1) Fisher’s exact test (p ≤ 0.005) for the duplication-only, deletion-only and U-shape models and Cochran-Armitage test (p ≤ 0.0005) for the mirror model; 2) linear regression (p ≤ 0.005) of the inverse normal quantile transformed trait residuals obtained from the logistic regression model of the binary outcome on the selected covariates.

### Enrichment analysis

For each gene, two groups of traits were defined: traits linked to the focal gene implicated by HPO *versus* other traits related to other genes in the 22q11.2 region but not to the focal gene. Association p-values for each probe within the gene (+/-10 kb) and each association model were compared between traits in the two groups using a one-sided Wilcoxon rank-sum test (i.e., H_a_: unrelated traits have lower association p-values with the focal gene than related ones). The number of effective tests (see **CNV probe selection and number of effective tests**) for each gene was calculated and used to define gene-specific significance thresholds. Genes were considered significant when the probe with the smallest p-value reached that threshold. The comparison was only performed for genes with at least four continuous traits and ten binary traits in each group. A binominal enrichment was performed to establish whether the number of genes significant in the Wilcoxon rank-sum test was higher than expected by chance with *pbinom* function in R.

### Transcriptome-wide Mendelian Randomization (TWMR)

TWMR was conducted as previously described ^18^ to identify changes in transcript levels of genes in the 22q11.2 region that causally modulate traits found to be associated to 22q11.2 CNVs by our association scan and, if this was the case, in which direction (i.e., whether increased gene expression associates with increased or decreased phenotype value). Briefly, the exposure (i.e., transcript level) and outcome (i.e., trait) are instrumented using independent genetic variants (instrumental variables (IVs); r^2^ < 0.01). Given their genetic effect sizes on these two quantities, a causal effect of the exposure on the outcome can be estimated using two-sample MR. Genetic effect sizes on transcript levels originate from whole blood expression quantitative trait loci (eQTL) provided by the eQTLGen consortium (cis-eQTLs at FDR < 0.05, 2-cohort filter) ^19^. Effect sizes on the traits stem from genome-wide association study (GWAS) summary statistics conducted on the UK Biobank (Neale’s lab: http://www.nealelab.is/uk-biobank/; Pan-UKBB team: https://pan.ukbb.broadinstitute.org) (**Table S6**). Prior to the analysis, eQTL and GWAS data were harmonized, palindromic SNPs were removed, as well as SNPs with an allele frequency difference > 0.05 between datasets. For increased robustness of the estimated causal effects, ≥ 5 (independent) IVs were required. MR estimates were considered significant when p ≤ 0.05/17 = 0.003 to account for the testing of 17 transcripts with at least 5 IVs and only significant genes overlapped by the CNV association signal were reported.

TWMR results were used for validation of the mirror model associations. It is expected that TWMR and mirror model effects are directionally concordant, i.e., increase/decrease in copy-number has the same direction of effect on a trait as an increase/decrease in gene expression. For this purpose, nominally significant (p < 0.05) TWMR effects were retained and their direction was compared to the direction of the probe with the smallest nominally significant p-value (p < 0.05) in the mirror association model for the corresponding gene (± 10 kb) and trait.

### Multivariable Mendelian Randomization (MVMR)

MVMR was performed to assess the causal relationship between significantly associated traits and compute a phenotype network. IVs were obtained from Neale Lab UKBB (http://www.nealelab.is/uk-biobank) and Pan-UKBB (https://pan.ukbb.broadinstitute.org) (**Table S6**) GWAS summary statistics for all eight significant continuous traits and nine significant binary traits. Data were harmonized with genetic variants in the UK10K reference dataset and variants with minor allele frequency (MAF) ≤ 0.01 were filtered out. Genetic variants were clumped at r^2^ = 0.001 using UK10K as a reference panel in PLINK v1.9. Mendelian randomization (MR) analysis was performed in two steps. First, potentially causal effects were identified with a univariable inverse variance weighted (IVW) MR for all exposure-outcome combinations (i.e. pairs of associated traits). Second, all exposures with nominally significant IVW causal effect estimates for a given outcome were included in an MVMR analysis as exposures. To reduce bias due to potential reverse causation, Steiger filtering was performed in all MR analyses (p < 5×10^−3^).

MVMR established the causal relationships among assessed traits using genetic variants as IVs. To infer if the pleiotropic effect of CNVs is vertical (indirect) or horizontal (genuine), we estimated what would be the expected CNV effect on the outcome trait (β_expected outcome_) if that outcome is a downstream result of the exposure trait as suggested by the MVMR analysis (vertical pleiotropy). β_expected outcome_ was determined as β_exposure_ × β_IVW_, with β_exposure_ the effect size of the best probe in the mirror model for each exposure (i.e. observed CNV - exposure trait association) and β_IVW_ the causal estimate for each exposure-outcome pair obtained from IVW MR. We then compared β_expected outcome_ with the observed CNV effect on the outcome trait (β_observed outcome_) obtained from the mirror association model.

### Software versions

Genetic analyses were conducted with PLINK v1.9 and PLINK v2.0. Statistical analyses were performed with R v3.6.1 and figures were generated with R v4.2.0.

## RESULTS

### 22q11.2 CNVs in the UKBB

After CNV calling and quality control in 405’324 unrelated individuals of the UKBB, we identified 1’127 individuals with a duplication and 694 individuals with a deletion overlapping the 22q11.2 LCRA-D region (**Figure 1A**). CNVs varied in size: duplication length ranged between 71 kb and 8.8 Mb (i.e., breakpoints extending beyond the defined region) with a median of 132 kb, while deletion length ranged between 80 kb and 2.8 Mb also with a median of 132 kb.

**Figure 1.**
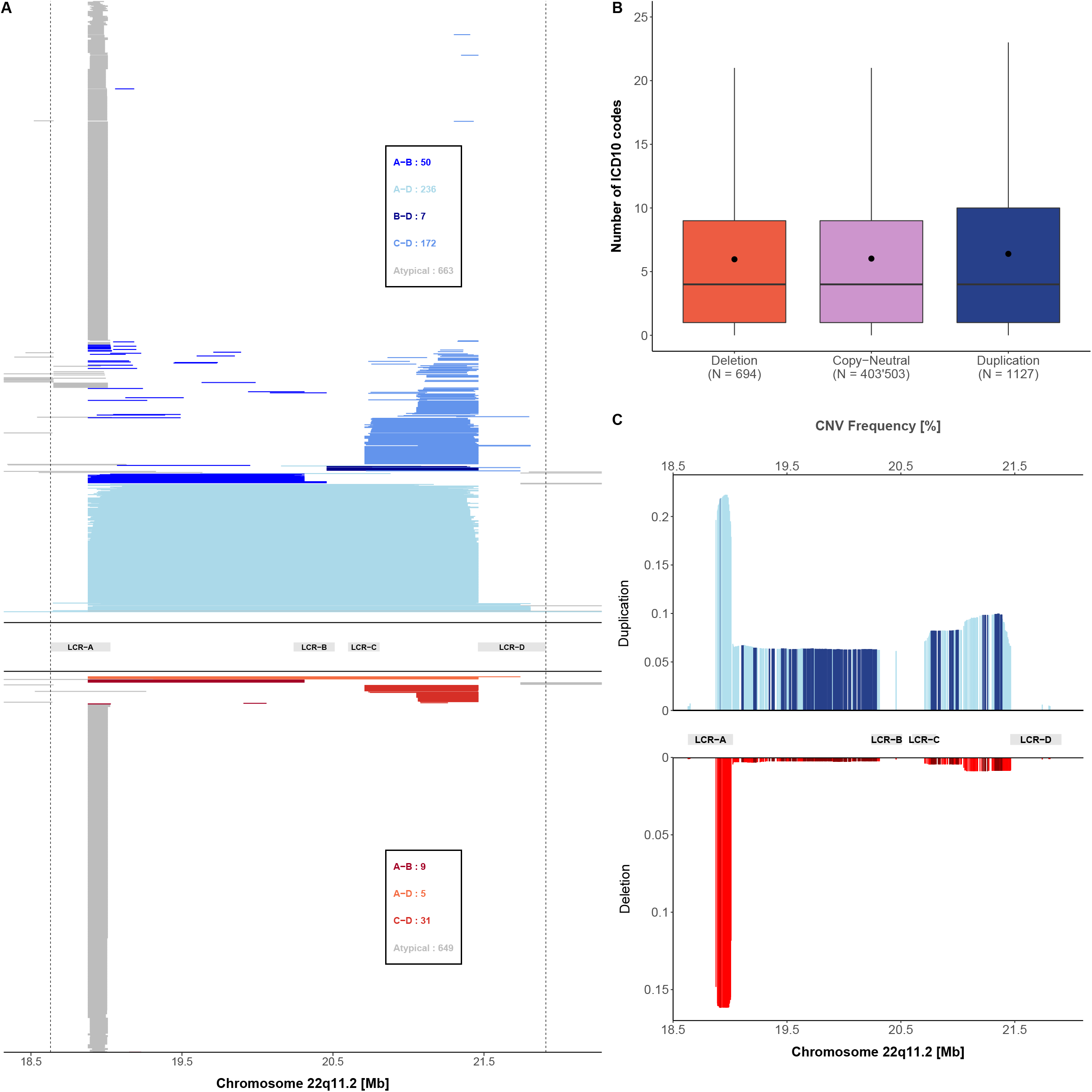
22q11.2 CNVs landscape. **A)** Each UKBB CNV carrier is displayed through a segment than spans the genomic coordinates of the CNV. Duplications are represented in the top part of the graph, while deletions at the bottom. Shades of blue and red represent different duplication and deletion categories, respectively, according to their localization in reference to the LCRA to LCRD. The number of duplications and deletions for each category is displayed in the boxes. **B)** Boxplot representing the number of ICD-10 codes reported in individuals grouped according to their copy-number state in the 22q11.2 region. N indicates the sample size for each category; dots show the mean; boxes show the first (Q1), second (median, thick line), and third (Q3) quartiles; lower and upper whiskers show the most extreme value within Q1 minus and Q3 plus 1.5× the interquartile range; outliers are not shown. **C)** Probe-level duplication (top, blue) and deletion (bottom, red) frequencies [%] for 864 probes plotted against the 22q11.2 genomic region. Frequency was calculated as the number of duplications or deletions divided by the total number of individuals assessed for the probe.

To assess whether individuals with these CNVs (mean number of diagnoses = 8.6) had a higher disease burden than individuals that are copy-neutral within this region (mean number of diagnoses = 8), we compared the reported number of ICD-10 codes and identified no statistical difference (two-sided Wilcoxon rank-sum test: p_del_= 0.44; p_dup_= 0.053) (**Figure 1B**).

CNVs were classified according to their localization as defined by LCRA-D. Between LCRs A and B, duplications were identified at a frequency of 0.01% and deletions at 0.002%; CNVs from LCR A to D, had a frequency of 0.06% and 0.001% for duplications and deletions respectively; from LCR B to D, duplications had a frequency of 0.002% and no deletions were identified; between LCRs C and D, duplications were identified at a frequency of 0.04% while deletions at 0.008%. CNVs that did not fall into these categories were considered as atypical and had a frequency of 0.16% for both duplications and deletions (**Figure 1A)**.

To account for all CNVs and bypass issues related to breakpoint variability, CNV calls were converted into probe-by-sample matrices for the CNV association scan. Probe-level CNV frequency after excluding LCRA probes (mean duplication frequency: 0.07%; mean deletion frequency: 0.004%) ranged between 0.004-0.1% and 0.001-0.01% for duplications and deletions, respectively **(Figure 1C**).

### Associated Traits

CNV association scan revealed significant links for eight continuous (**Table 2, Figure S1**) and nine binary traits (**Table 3, Figure S2**), which were associated under different association models. Eight traits (four binary and four continuous) were associated most significantly under the U-shape model, three continuous traits did so under the mirror model, four binary traits were associated more significantly under the duplication-only model and two traits under the deletion-only model (one continuous and one binary), highlighting the importance of testing models mimicking different dosage mechanisms. Among the identified continuous traits, body mass index (BMI) was found associated under the U-shape model (β = 1.56 kg/m^2^, p = 4.9 × 10^−10^) throughout LCRA to LCRD (**Figure 2A**) indicating that both duplications and deletions increase BMI level (**Figure 2B**). TWMR analysis showed that increased expression of *ARVCF* increases BMI (β = 0.05, p = 10^−4^), concordantly with the positive association found by the mirror CNV association scan (**Figure 2C**).

**Figure 2.**
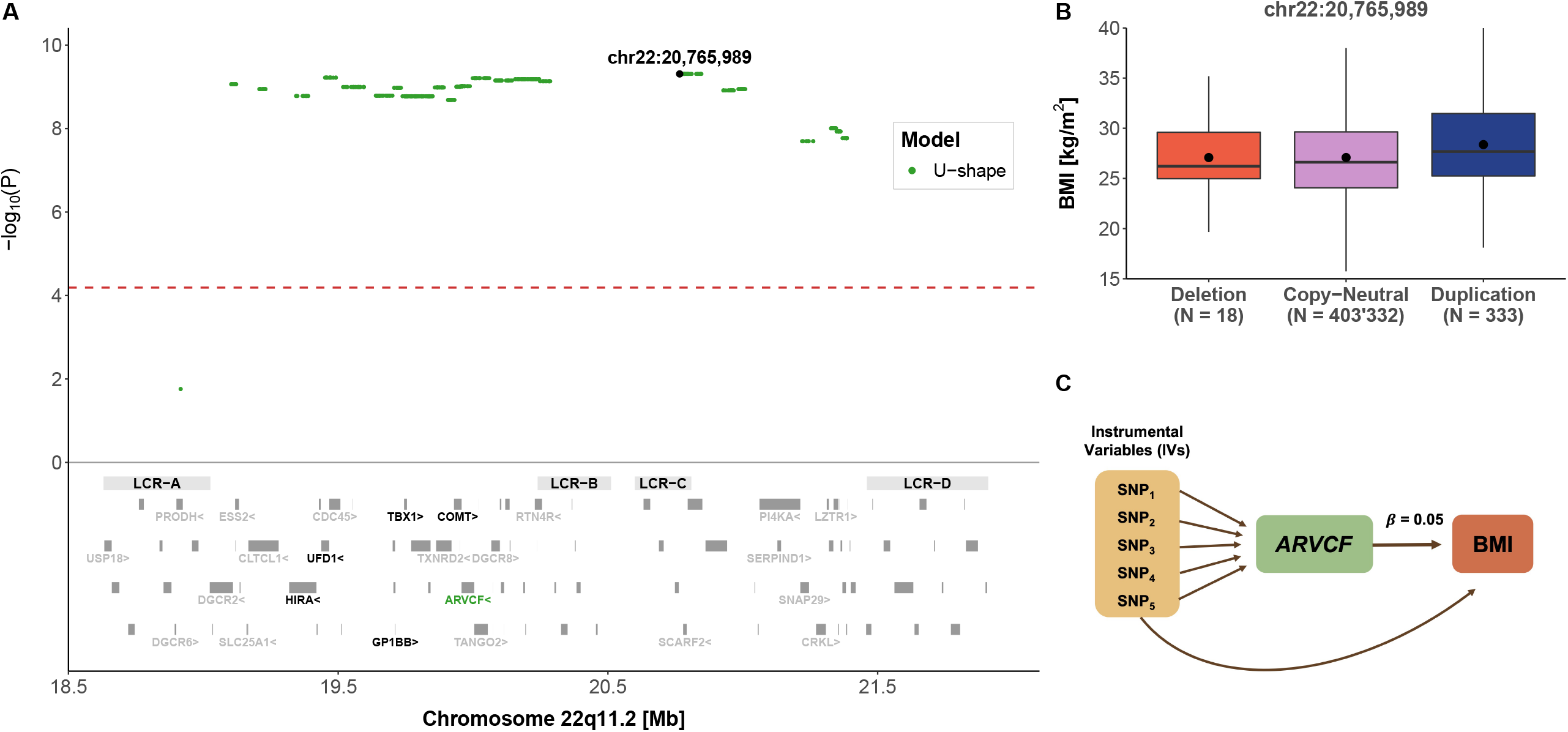
22q11.2 CNVs and body mass index (BMI). **A) Top:** The negative logarithm of the association p-value for the U-shape CNV-BMI association scan is plotted against the 22q11.2 genomic region. Each point represents a CNV proxy probe and the lead signal (chr22:20,765,989) is shown in black. The red dashed line indicates significance threshold (p < 6.5 × 10^−5^). **Bottom:** Low copy-repeat region (LCR) A-D, as well as the 90 genes contained in the region. The 24 genes linked to traits according to HPO are labeled and genes linked to BMI through HPO are labeled in black. *ARVCF* expression was found to causally influence BMI through TWMR and is shown in green. **B)** Boxplot representing BMI in individuals grouped according to their copy-number state of the lead signal probe (chr22:20,765,989). N indicates the sample size for each category; dots show the mean; boxes show the first (Q1), second (median, thick line), and third (Q3) quartiles; lower and upper whiskers show the most extreme value within Q1 minus and Q3 plus 1.5× the interquartile range; outliers are not shown. **C)** Representation of the TWMR analysis showing SNPs as instrumental variables (IVs), *ARVCF* gene expression as exposure, and its causal effect size (β = 0.05) on BMI.

**Table 2.** Continuous traits associated to CNVs in the 22q11.2 region with different models.

| Phenotype | Genomic Position | Duplication-only |  |  | Deletion-only |  |  | U-shape |  |  | Mirror |  |  |
| --- | --- | --- | --- | --- | --- | --- | --- | --- | --- | --- | --- | --- | --- |
| | | $\beta$ | 95 % CI | p-value | $\beta$ | 95 % CI | p-value | $\beta$ | 95 % CI | p-value | $\beta$ | 95 % CI | p-value |
| Mean Platelet Volume (femtolitres) | chr22:19639383 | -0.54 | [-0.67,-0.41] | $1.16 \times 10^{-15}$ | 1.66 | [0.99,2.32] | $1.13 \times 10^{-6}$ | -0.46 | [-0.59,-0.33] | $4.97 \times 10^{-12}$ | **-0.58** | **[-0.71,-0.45]** | **$1.31 \times 10^{-18}$** |
| Body mass index (Kg/m<sup>2</sup>) | chr22:20765989 | 1.65 | [1.15,2.16] | $1.55 \times 10^{-10}$ | -0.06 | [-2.23,2.12] | 0.96 | **1.56** | **[1.07,2.06]** | **$4.9 \times 10^{-10}$** | 1.57 | [1.08,2.06] | $4.23 \times 10^{-10}$ |
| Whole body fat mass (kg) | chr22:20765989 | 3.17 | [2.18,4.16] | $3.70 \times 10^{-10}$ | -1.74 | [-6.37,2.88] | 0.46 | 2.95 | [1.98,3.92] | $2.33 \times 10^{-9}$ | **3.11** | **[2.14,4.07]** | **$3.35 \times 10^{-10}$** |
| Fluid intelligence score | chr22:19343881 | -1.21 | [-1.64,-0.79] | $2.25 \times 10^{-8}$ | -3.76 | [-6.04,-1.49] | 0.001 | **-1.3** | **[-1.72,-0.88]** | **$1.12 \times 10^{-9}$** | -1.04 | [-1.46,-0.63] | $9.54 \times 10^{-7}$ |
| Weight (kg) | chr22:20765989 | 3.83 | [2.33,5.32] | $5.63 \times 10^{-7}$ | -4.28 | [-11.28,2.73] | 0.231 | 3.47 | [2.01,4.94] | $3.44 \times 10^{-6}$ | **3.85** | **[2.38,5.31]** | **$2.70 \times 10^{-7}$** |
| Height (cm) | chr22:21219710 | -0.6 | [-1.23,0.03] | 0.064 | **-4.86** | **[-6.96,-2.77]** | **$5.51 \times 10^{-6}$** | -0.95 | [-1.56,-0.35] | 0.002 | -0.14 | [-0.75,0.46] | 0.64 |
| Height (cm) | chr22:19518079 | -1.94 | [-2.72,-1.15] | $1.43 \times 10^{-6}$ | -6.02 | [-10,-2.04] | 0.003 | **-2.09** | **[-2.86,-1.32]** | **$1.14 \times 10^{-7}$** | -1.64 | [-2.41,-0.86] | $3.26 \times 10^{-5}$ |
| Platelet Count (10<sup>9</sup> cells/L) | chr22:19738355 | 16.68 | [9.56,23.8] | $4.43 \times 10^{-6}$ | -100.09 | [-135.83,-64.35] | $4.05 \times 10^{-8}$ | 12.22 | [5.24,19.21] | 0.0006 | **19.86** | **[12.88,26.85]** | **$2.48 \times 10^{-8}$** |
| Calcium level (mmol/L) | chr22:19207491 | 0.01 | [0,0.02] | 0.089 | **-0.13** | **[-0.18,-0.08]** | **$2.86 \times 10^{-7}$** | 0.003 | [-0.01,0.01] | 0.64 | 0.02 | [0.01,0.03] | 0.004 |

Reported effect sizes and p-values for each model are referring to the lead signal of the most relevant model for each phenotype (in bold).

**Table 3.** Binary traits associated to CNVs in the 22q11.2 region with different models.

| Phenotype | Genomic Position | Duplication-only |  |  | Deletion-only |  |  | U-shape |  |  | Mirror |  |  |
| --- | --- | --- | --- | --- | --- | --- | --- | --- | --- | --- | --- | --- | --- |
|  |  | OR | 95 % CI | p-value | OR | 95 % CI | p-value | OR | 95 % CI | p-value | OR | 95 % CI | p-value |
| Gastroesophageal reflux disease | chr22:19998655 | **2.72** | **[1.91,3.88]** | **$2.53 \times 10^{-8}$** | 1.65 | [0.21,13.01] | 0.63 | 2.68 | [1.89,3.79] | $2.69 \times 10^{-8}$ | 2.66 | [1.87,3.79] | $6.23 \times 10^{-8}$ |
| Hearing loss | chr22:20082293 | 4.47 | [2.49,8.02] | $5.32 \times 10^{-7}$ | 12.9 | [1.58,105.24] | 0.017 | **4.71** | **[2.68,8.27]** | **$6.95 \times 10^{-8}$** | 4.08 | [2.22,7.5] | $5.87 \times 10^{-6}$ |
| Cardiomegaly | chr22:21370246 | **3.53** | **[1.92,6.47]** | **$4.69 \times 10^{-5}$** | 4.78 | [0.64,35.95] | 0.13 | 3.6 | [2.02,6.45] | $1.53 \times 10^{-5}$ | 3.21 | [1.71,6.03] | 0.0003 |
| Dental caries | chr22:21370246 | 3.29 | [1.85,5.85] | $5.21 \times 10^{-5}$ | 5.94 | [1.4,25.12] | 0.015 | **3.51** | **[2.06,5.99]** | **$4.21 \times 10^{-6}$** | 2.76 | [1.48,5.13] | 0.001 |
| Diplopia and disorders of binocular vision | chr22:21219710 | **6.23** | **[2.57,15.09]** | **$5.18 \times 10^{-5}$** | 7.31 | [0.43,124.7] | 0.17 | 5.74 | [2.37,13.92] | 0.0001 | 6.24 | [2.58,15.1] | $4.89 \times 10^{-5}$ |
| Other venous embolism and thrombosis | chr22:20765989 | **7.6** | **[2.82,20.46]** | **$6 \times 10^{-5}$** | 18.57 | [1.01,340.01] | 0.049 | 7.24 | [2.69,19.49] | $8.9 \times 10^{-5}$ | 7.61 | [2.83,20.46] | $5.86 \times 10^{-5}$ |
| Other cerebral degenerations | chr22:20927716 | 1.76 | [0.44,7.07] | 0.428 | **45** | **[10.05,201.43]** | **$6.43 \times 10^{-7}$** | 3.38 | [1.26,9.1] | 0.016 | 0.21 | [0.01,3.57] | 0.28 |
| Hypotension | chr22:20927716 | 3.16 | [1.79,5.6] | $7.7 \times 10^{-5}$ | 7.39 | [0.89,61.65] | 0.065 | **3.3** | **[1.9,5.73]** | **$2.16 \times 10^{-5}$** | 2.91 | [1.61,5.25] | 0.0004 |
| Nausea and vomiting | chr22:21370246 | 2.17 | [1.42,3.31] | 0.0003 | 3.67 | [1.09,12.37] | 0.036 | **2.28** | **[1.53,3.39]** | **$5.34 \times 10^{-5}$** | 1.92 | [1.24,2.98] | 0.003 |

Reported OR and p-values for each model are referring to the lead signal of the most relevant model for each phenotype (in bold)

Mean platelet volume (MPV) was found associated under the mirror model (β = -0.58 femtolitres, p = 1.3 × 10^−18^) with the strongest association occurring in the LCRA to LCRB region (**Figure 3A**). The signal replicated in both the duplication-only (β = -0.54 femtolitres, p = 1.16 × 10^−15^) and deletion-only (β = 1.66 femtolitres, p = 1.13 × 10^−6^) model providing further evidence of a “true mirror” effect, despite the deletion effect being slightly stronger than the duplication one (**Figure 3B**). In line with this effect, TWMR revealed that increased *DGCR6* expression causally reduces MPV (β = -0.03, p = 0.001) (**Figure 3C**). It is worth noting that this trait is negatively correlated with platelet count (also significant under the mirror model, β = 19.86 10^9^ cells/L, p = 2.5 × 10^−8^). As expected, MVMR showed bidirectional causality between both traits, highlighting the challenges on interpreting their association separately.Unlike other phenotypes, height was associated under different models in distinct regions.

**Figure 3.**
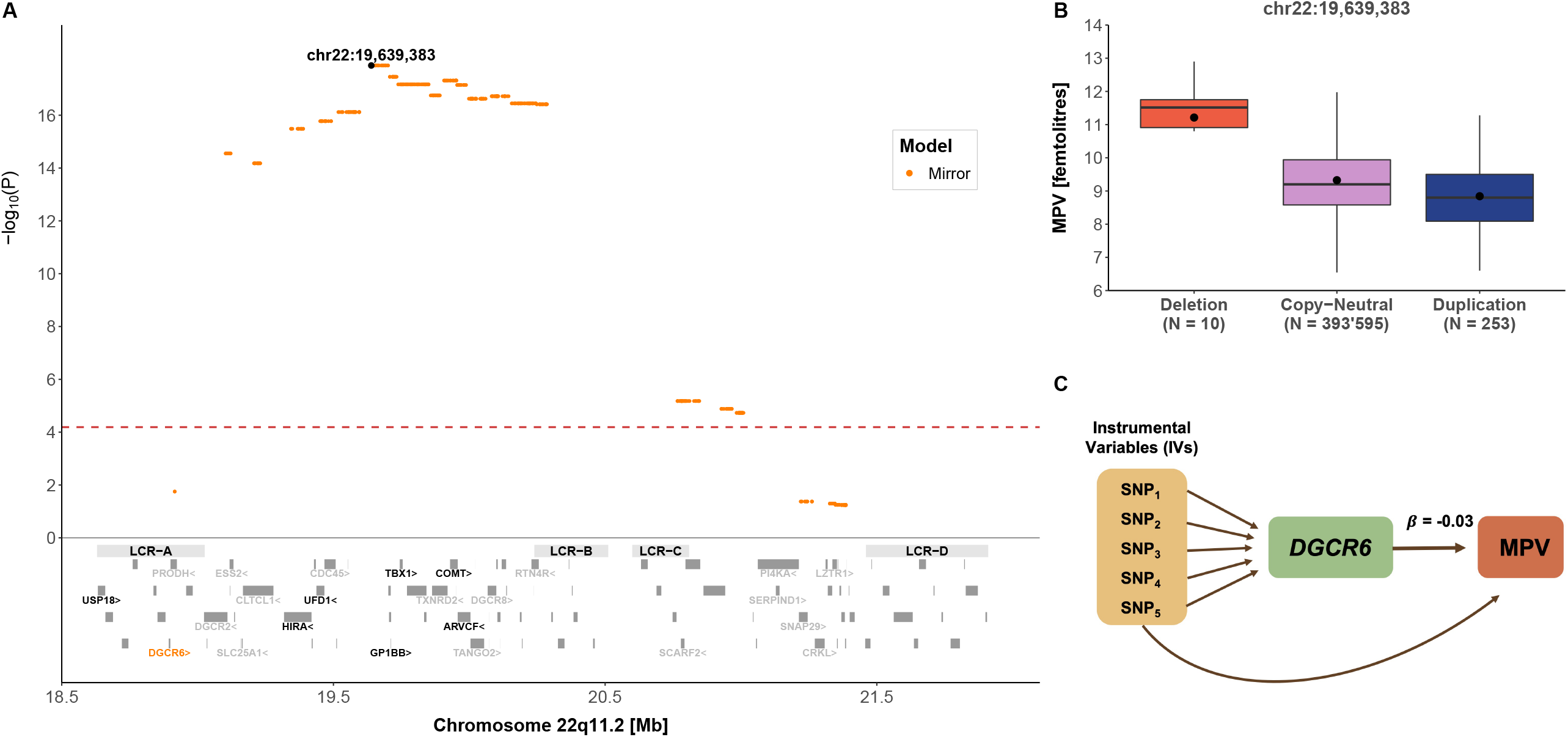
22q11.2 CNVs and mean platelet volume (MPV). **A) Top:** The negative logarithm of the mirror association p-value for the CNV-MPV association is plotted against the 22q11.2 genomic region. Each point represents a CNV proxy probe and the lead signal (chr22:19,639,383) is shown in black. The red dashed line indicates significance threshold (p < 6.5 × 10^−5^). **Bottom:** Low copy-repeat region (LCR) A-D, as well as the 90 genes contained in the region. The 24 genes linked to traits according to HPO are labeled and genes linked to mean platelet volume through HPO are labeled in black. *DGCR6* expression was found to causally influence mean platelet volume through TWMR and is shown in orange. **B)** Boxplot representing mean platelet volume in individuals grouped according to their copy-number state for the lead signal probe (chr22:19,639,383). N indicates the sample size for each category; dots show the mean; boxes show the first (Q1), second (median, thick line), and third (Q3) quartiles; lower and upper whiskers show the most extreme value within Q1 minus and Q3 plus 1.5× the interquartile range; outliers are not shown. **C)** Representation of the TWMR analysis showing SNPs as instrumental variables (IVs), DGCR6 gene and its causal effect size (β = - 0.03) on MPV.

The U-shape model appeared as the most significant model in the region spanning LCRA to LCRB (β = -2.09 cm, p = 1.1 × 10^−7^), while the deletion-only model was the only significant one at the distal portion between LCRC and LCRD (β = -4.86 cm, p = 5.5 × 10^−6^) (**Figure 4A**). Given this unexpected pattern, we stratified CNVs according to LCR categories (**Figure 1A**) to inspect their impact on height. Within LCRA-LCRB and LCRA-LCRD (**Figures 4B and 4C**), both duplications and deletions were associated with a height decrease in concordance with the U-shape model. However, duplications and deletions within LCRC and LCRD had opposing effects on height, in line with a mirror model which was confirmed by linear regression (β= 0.17 cm, p = 0.0003) (**Figure 4D**).

**Figure 4.**
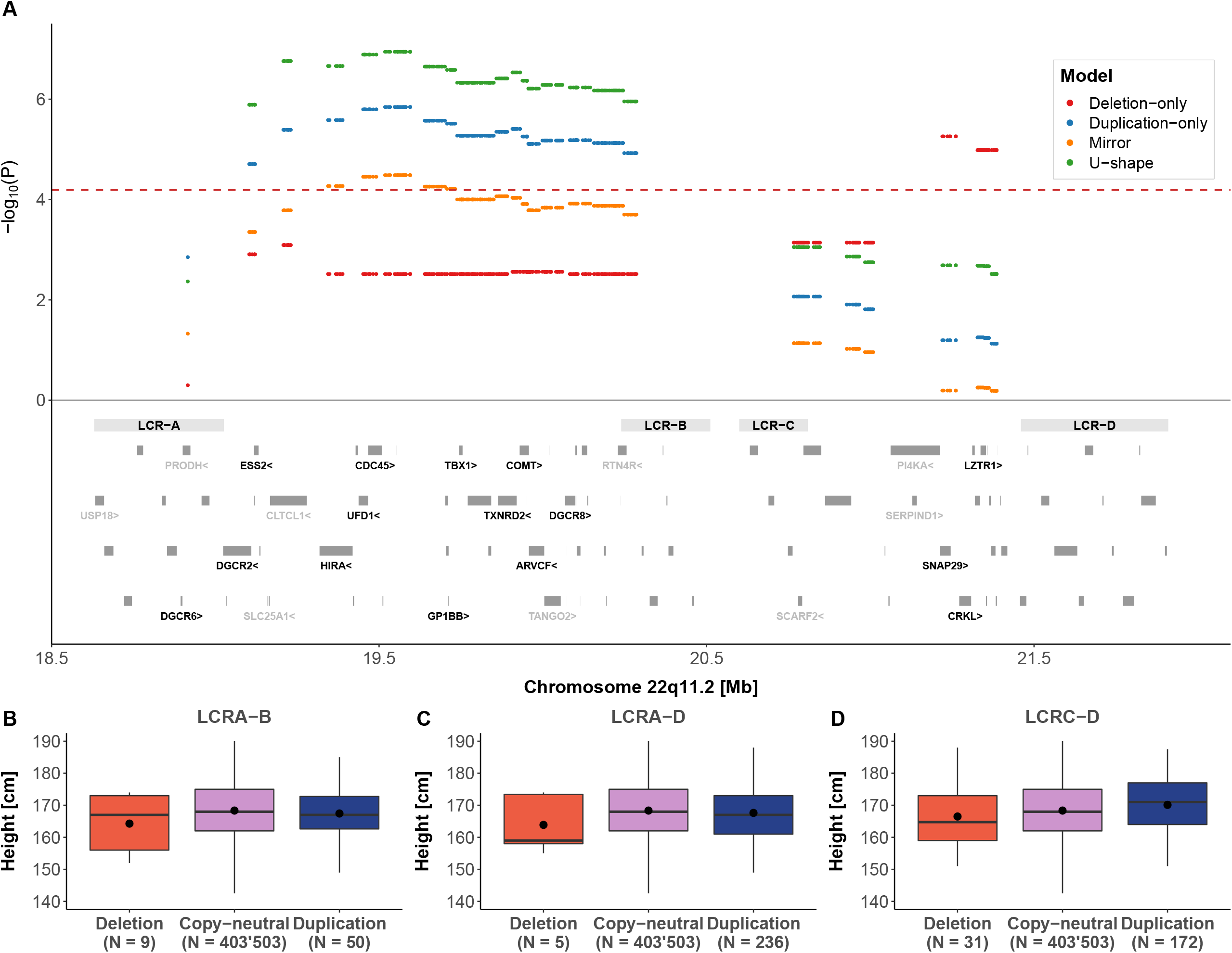
22q11.2 CNVs and height. **A) Top:** The negative logarithm of the association p-value for the CNV-height association according to a deletion-only (red), duplication-only (blue), mirror (orange), and U-shape (green) is plotted against the 22q11.2 genomic region. The red dashed line indicates significance threshold (p < 6.5 × 10^−5^). **Bottom:** Low copy-repeat region (LCR) A-D, as well as the 90 genes contained in the region. The 24 genes linked to traits according to HPO are labeled and genes linked to height through HPO are labeled in black. **B)** Boxplots representing height in individuals with **(B)** LCRA-B, **(C)** LCRA-D, and **(D)** LCRC-D CNVs grouped according to their copy-number state. N indicates the sample size for each category; dots show the mean; boxes show the first (Q1), second (median, thick line), and third (Q3) quartiles; lower and upper whiskers show the most extreme value within Q1 minus and Q3 plus 1.5× the interquartile range; outliers are not shown.

Given the low number of deletion carriers affected by binary outcomes (0 to 3 carriers) (**Table S7**), associations found under the U-shape or mirror models often reflect the effect of duplications (i.e., the most common CNV type) in these phenotypes. One example is gastroesophageal reflux disease which was found to be associated under the duplication-only model (OR = 2.72, p = 2.53 × 10^−8^) with a stronger association occurring in the LCRA to LCRB region (**Figure 5A**), indicating an increased prevalence of gastroesophageal reflux disease among duplication carriers (**Figure 5B**).

**Figure 5.**
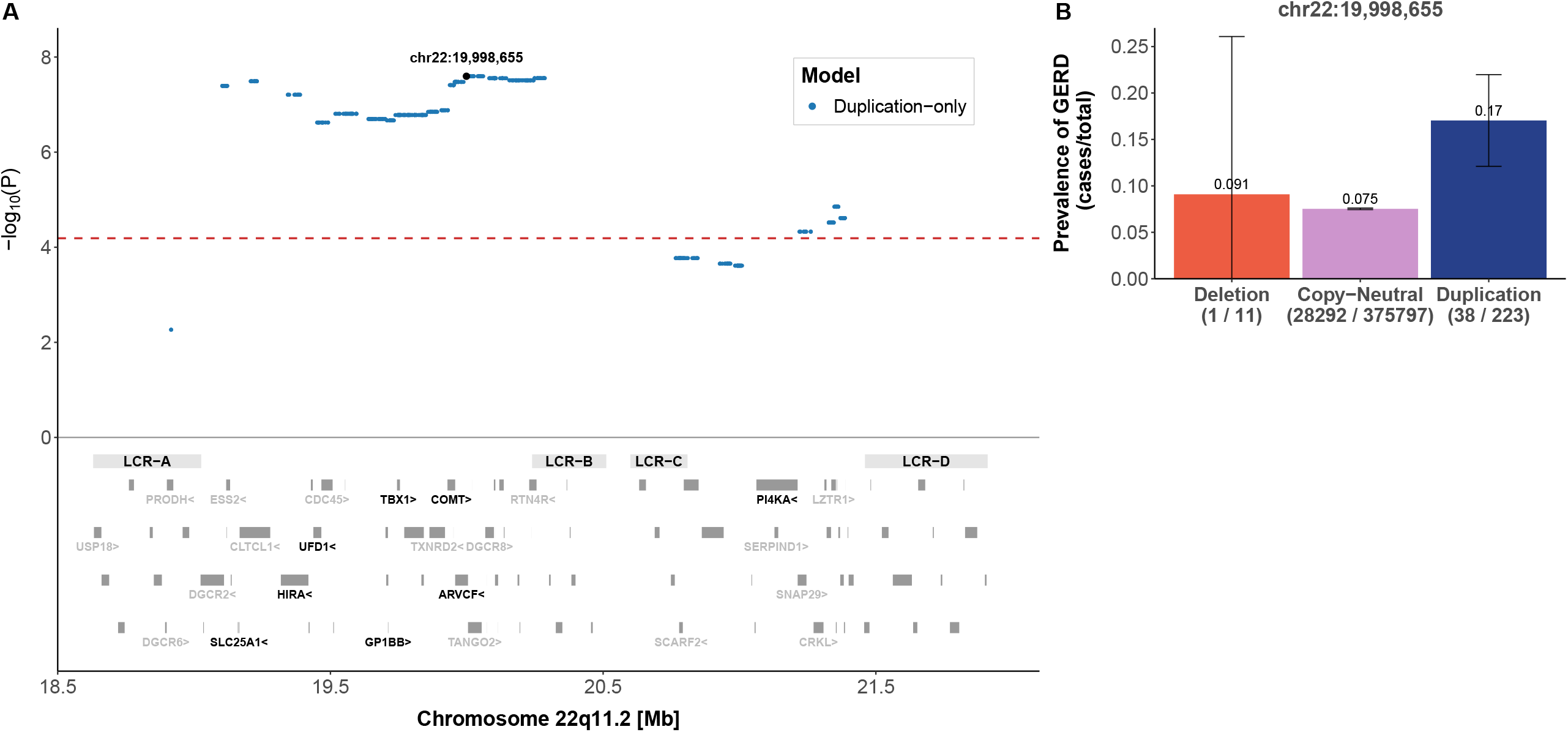
22q11.2 CNVs and gastroesophageal reflux disease (GERD). **A) Top:** The negative logarithm of the duplication-only association p-value for the CNV-GERD association is plotted against the 22q11.2 genomic region. Each point represents a CNV proxy probe and the lead signal (chr22:19,998,655). The red dashed line indicates significance threshold (p < 6.5 × 10^−5^). **Bottom:** Low copy-repeat region (LCR) A-D, as well as the 90 genes contained in the region. The 24 genes linked to traits according to HPO are labeled and genes linked to mean platelet volume through HPO are labeled in black. **B)** Barplot representing prevalence (cases/total) of GERD grouped according to copy-number state for the lead signal probe (chr22:19,998,655). 95% confidence interval for deletion is truncated at zero.

### Enrichment analysis

For continuous traits, 6 out of 8 assessed genes were found to have significantly greater association p-values for the group of unrelated traits compared to the group of linked traits for all association models (see Method section: Enrichment analysis for the definition of these groups). Binomial enrichment analysis indicated that CNV probes in genes linked to a given HPO term are 15 times more likely (p < 6 ×10^−9^) to show stronger association with the corresponding UKBB continuous trait. For the binary traits, however, only 2 out of 19 assessed genes were significant in the mirror model which does not indicate an enrichment (p = 0.07).

### Concordance in the direction of effect between association scan and TWMR

Besides showing that differential expression of two 22q11.2 genes (*ARVCF* and *DGCR6*) causally affects two associated traits (BMI and MPV), TWMR results were also used to reinforce reliability of CNV associations. We evaluated concordance in the direction of effect sizes from nominally significant (p < 0.05) results of the mirror CNV association scan and nominally significant (p < 0.05) TWMR results (**Table S8**). As expected, we observed a significant agreement in effect size directions between both when fitting a linear regression line (β = 1.6, p = 0.01; **Figure 6**).

**Figure 6.**
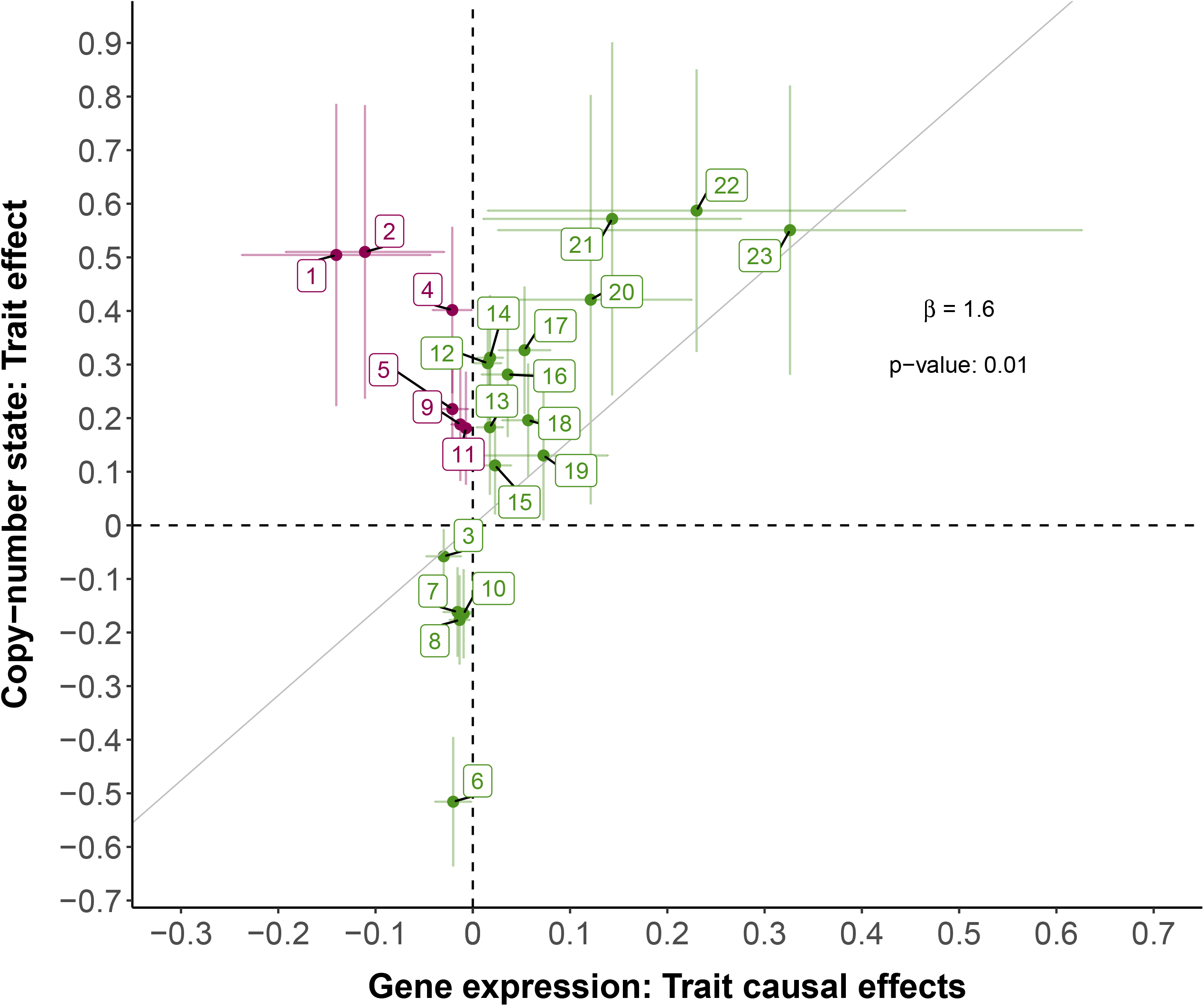
Concordance between TWMR and CNV association scan effect sizes: Scatter plot depicting mirror association scan (y-axis) versus TWMR (x-axis) effect sizes. Vertical and horizontal bars represent the 95% confidence intervals. The (zero-intercept) regression line and the corresponding slope are in black. For association scan effect sizes, the probe with the smallest p-value in the mirror model located in the TWMR gene was selected. Trait-gene pairs with agreeing direction between TWMR and CNV association scan are in green and trait-gene pairs with opposite directions are in pink. Labels indicate: (1) hypotension - *GNB1L*; (2) cardiomegaly - *P2RX6*; (3) mean platelet volume - *DGCR6*; (4) gastroesophageal reflux disease - *GNB1L*; (5) weight - *P2RX6*; (6) mean platelet volume - *CLDN5*; (7) height - *TANGO2*; (8) height - *CLDN5*; (9) weight - *CLDN5*; (10) height - *GNB1L*; (11) weight - *GNB1L*; (12) body mass index - *SLC25A1*; (13) calcium levels - *CLTCL1*; (14) platelet count - *CLDN5*; (15) platelet count - *P2RX6*; (16) whole body fat mass – *ARVCF*; (17) body mass index – *ARVCF*; (18) weight – *ARVCF*; (19) nausea and vomiting - *DGCR6*; (20) diplopia and disorders of binocular vision - *DGCR6*; (21) cardiomegaly – *ARVCF*; (22) hearing loss - *SLC25A1*; (23) hypotension - *DGCR2*.

### Causal links between traits and CNV pleiotropy

Cross-trait MVMR was performed for all 17 significantly associated traits. Out of a total of 289 trait-pair combinations, we identified 48 pairs that are causally linked to each other at nominal significance (p < 0.05) using the IVW MR method. MVMR was then applied on these 48 combinations and 17 trait-pairs were significant after Bonferroni correction (p < 0.05/289 = 0.0002) (**Figure 7A**). Most traits were associated in a bidirectional manner, indicating that many (closely related) traits are mutually related to each other likely due to high genetic correlation. To distinguish between horizontal and vertical pleiotropy, we plotted the CNV effect on the outcome expected under vertical pleiotropy (β_expected outcome_) against the effect observed in the association scan (β_observed outcome_) to examine the concordance in effect direction (**Figure 7B;** MVMR method section). This analysis revealed agreement only for very closely related trait pairs (driven by strong genetic correlation) such as platelet count – mean platelet volume, and indicated that, in general, pleiotropic CNV association are not due to vertical, but rather due to genuine horizontal pleiotropy.

**Figure 7.**
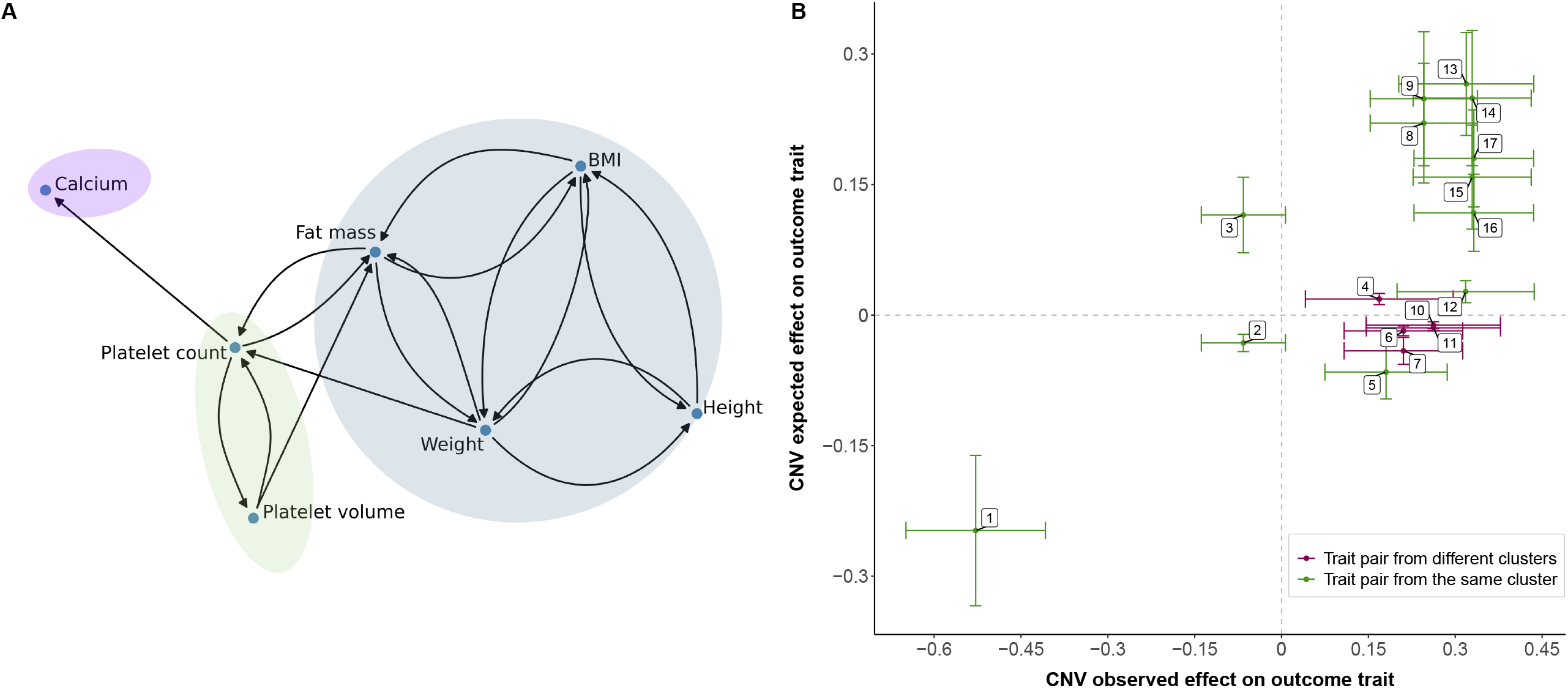
Concordance between CNV expected and observed effect on outcome trait: **(A)** Causal links identified in the MVMR analysis. Colored shapes indicate clusters of traits grouped based on their correlation (r > |0.45|). **(B)** Scatter plot depicting estimated CNV expected effect on the outcome (y-axis) versus CNV observed effect on outcome (x-axis) for each trait pair. Trait pairs from the same cluster (**A**) are in green and trait pairs from different clusters are in pink. The vertical and horizontal bars represent the 95% confidence intervals. Labels indicate exposure – outcome pairs: (1) platelet count – mean platelet volume; (2) body mass index – height; (3) weight – height; (4) platelet count - calcium levels; (5) height – weight; (6) fat mass - platelet count; (7) weight - platelet count; (8) body mass index – weight; (9) fat mass – weight; (10) platelet count - fat mass; (11) mean platelet volume - fat mass; (12) height – body mass index; (13) mean platelet volume - platelet count; (14) BMI - fat mass; (15) weight - fat mass; (16) weight – body mass index; (17) fat mass – body mass index.

## DISCUSSION

Most of our knowledge regarding the impact of CNVs in the 22q11.2 region in the general population stems from genome-wide studies ^10,20–24^. Here, we focused on this region specifically and developed a tailored set of analyses with more lenient, yet appropriate, significance threshold and in-depth follow-up analyses that allowed to detect plausible associations missed by genome-wide studies (e.g., hearing loss, cardiomegaly, diplopia, and disorders of binocular vision). Our findings show that 22q11.2 CNV carriers in the general population, that are likely on the milder end of the phenotypic spectrum, are associated with traits previously implicated by genes in the region, shedding light on the variable expressivity and penetrance of CNVs impacting this complex genomic region.

Assessed traits linked to 22q11.2 genes have been previously identified in different contexts including the 22q11.2 deletion and duplication syndromes, clinical conditions caused by variants in a single gene, and complex conditions associated with the locus (**Figure S3**). Therefore, using the HPO database to select investigated traits allowed us to leverage information from different genetic variants in a clinical context ^13^, to identify associations in the general population. Our enrichment analysis showed that for continuous traits this was an effective approach. We also show that CNVs can impact traits previously known to be associated with individual genes in the region, such as cardiomegaly (*LZTR1*, OMIM:616564) and other venous embolism and thrombosis (*SERPIND1*, OMIM:612356), that were both associated under the duplication-model in the distal region between LCRC and LCRD, which harbors these genes.

Our results validated several known associations and furthermore shed light on traits that have not yet been extensively studied in the context of 22q11.2 CNVs. For instance, gastroesophageal reflux disease is not a vastly explored clinical feature in 22q11.2 deletion or duplication syndromes. While LCRA to LCRD duplications have been previously associated with this trait in the UKBB cohort ^20^, replication of the association in our study emphasizes its relevance in 22q11.2 CNV carriers. Another relevant association identified in our study is with BMI. Obesity (BMI > 30) is a well-known phenotype in individuals with 22q11.2DS ^25^. Even though this phenotype is not well described in clinical studies characterizing the 22q11.2 duplication syndrome, an increase in BMI has been associated with duplications in other studies assessing the UKBB cohort ^10,21^. We have further shown a causal effect of differential expression of *ARVCF* – a gene whose product is part of the catenin family and is involved in protein-protein interactions at adherent junctions (OMIM: 602269) – on BMI. Recently, a rare *ARVCF* missense variant of unknown significance has been identified in an individual with early-onset severe obesity ^26^, suggesting that *ARVCF* may play an important role in the etiology of obesity.

Besides validating the link between CNVs in the 22q11.2 region and platelet count ^10^, we revealed a new association with mean platelet volume which exhibits a “true mirror” effect, reinforcing the role of this genomic region in phenotypes such as thrombocytopenia. Thrombocytopenia is a well-known clinical hallmark in 22q11.2DS ^1^ but is not yet recognized as a clinical feature of the 22q11.2 duplication syndrome. *GP1BB* represents a top candidate to explain the observed platelet phenotypes as biallelic loss of function variants in the gene are responsible for Bernard-Soulier Syndrome, a platelet disorder (OMIM: 231200), and inclusion of *GP1BB* in the deleted region has been implicated in reduction of platelet count levels in 22q11.2DS patients ^27^. Due to lack of sufficient IVs, *GP1BB* could not be assessed by TWMR analysis, which instead revealed a causal effect of *DGCR6* differential expression on MPV. While *DGCR6*’s function is not yet clearly defined (OMIM: 601279), it has been implicated in regulating other genes in the 22q11.2 region ^28^, suggesting that multiple genes in the region influence platelet phenotypes.

Usage of four different association models allowed for the identification of deletion-specific effects (e.g., calcium level), as well as traits in which duplications and deletions act in the same or in opposite directions. By performing association scans at the probe level, we also showed that gain or loss of distinct segments within 22q11.2 may impact a trait following different association models, as was seen for height. Short stature has been identified for the 22q11.2DS ^1^ but variable height measures have been described for the 22q11.1 duplication syndrome ^29–31^. In concordance with our study, both duplications and deletions (LCRA to LCRD) have been previously associated with a decrease in height in the UKBB cohort ^21^. However, our study is the first to show a mirror behavior involving the LCRC to LCRD region. The impact of CNVs in the LCRC-D region is often overlooked or considered in combination with LCRA to LCRB. However, the unexpectedly distinct impact of CNVs in this region on height, as well as certain traits that were only significant in this region (such as weight, cardiomegaly, other venous embolism and thrombosis, dental caries), reveal the value of a more refined study of CNVs overlapping this complex region.

A drawback of studying pathogenic CNVs in a general population such as the UKBB is that the number of affected participants is low as carriers of 22q11.2 CNVs with larger phenotypic impact are less likely to participate, a phenomenon often described as the “healthy volunteer” selection bias ^32^. As such, frequencies of the 22q11.2 deletions and duplications have not been precisely estimated outside of clinical cohorts. In the general population, frequency of deletions and duplications encompassing the LCRA to LCRB region have been estimated at 0.02% and 0.08%, respectively (Kirov et al. 2014). Another study estimated a frequency of 0.03% for deletions and 0.07% for duplications considering the typical 3 Mb and 1.5 Mb CNVs ^34^. In our work, the frequency of CNVs in LCRA to LCRB and LCRA to LCRD is 0.07% for duplications and 0.003% for deletions. It is worth noting that we consider smaller nested CNVs between LCRA and LCRB that were not appreciated in previous studies, indicating that if we applied similar definitions to these works, our frequency estimates would be lower.

While the absolute number of CNV carriers considered in our study is still larger than the sample size of some clinical cohorts, these individuals tend to exhibit milder phenotypes. This hampers statistical power to detect associations, especially for binary outcomes for which trait definition through grouping of ICD-10 codes is imperfect and arbitrary and case number can be extremely low. We offer corroborating evidence of our findings’ reliability by performing sensitivity analyses and examining the concordance of CNV findings with TWMR effects. Importantly, effects observed in our study are potentially smaller than the ones observed in clinical cohorts ^35^ as they are mainly derived from CNV carriers with sub-clinical phenotypes and thus represent lower bound estimates. While in theory estimates from clinical cohorts might offer upper bound estimates, their poor and unstandardized reporting makes it difficult to establish accurate comparisons. Still, we hope that our study offers a better understanding on the spectrum of phenotypic consequences exerted by 22q11.2 and will improve diagnostic rates in individuals with low expressed phenotypes as molecular diagnostic of genomic syndromes still often relies on recognition of characteristic signs to guide genetic testing.

## CONCLUSION

We found that 22q11.2 CNVs affect traits compatible with clinical manifestations seen in the genomic disorders within the general population. The probe-level association scan revealed that dosage of different segments within the 22q11.2 region may impact a trait through different mechanisms, as illustrated with height. Besides, yielding further insights into the complex 22q11.2 region, our study provides a framework that can be adapted to study the phenotypic consequences of other clinically relevant genomic regions.

## Data Availability

All data produced in the present study are available upon reasonable request to the authors.

## ACKNOWLEDGMENTS

This study was conducted with the UK Biobank Resource (under application number 16389), we thank all biobank participants for sharing their data. This work was supported by funding from the Department of Computational Biology (Z.K.), the Swiss National Science Foundation (310030-189147) as well as financial support from Fundação de Amparo à Pesquisa do Estado de São Paulo [2020/11241-2, M.Z.; 2019/21644-0, M.I.M] and from the Coordenação de Aperfeiçoamento de Pessoal de Nível Superior [Brasil (CAPES).

## AUTHOR CONTRIBUTIONS

M.Z. contributed to study design, conducted analysis, and interpretation of the data and wrote the article. C.A. contributed to study design and interpretation of the data. M.C.S performed TWMR analysis. A.G. performed MVMR analysis. K.L. designed the web scraping approach used for mapping of HPO terms to UKBB traits. M.M.O., A.G.D. and M.I.M. contributed to study design and interpretation. Z.K. supervised the study, contributed to study design and interpretation of the data. All authors critically revised the manuscript and approved the final version.

## REFERENCES

1. McDonald-McGinn, D.M., Sullivan, K.E., Marino, B., Philip, N., Swillen, A., Vorstman, J.A.S., Zackai, E.H., Emanuel, B.S., Vermeesch, J.R., Morrow, B.E., et al. (2015). 22q11.2 deletion syndrome. Nat Rev Dis Primers 15071.

2. Monteiro, F.P., Vieira, T.P., Sgardioli, I.C., Molck, M.C., Damiano, A.P., Souza, J., Monlleó, I.L., Fontes, M.I.B., Fett-Conte, A.C., Félix, T.M., et al. (2013). Defining new guidelines for screening the 22q11.2 deletion based on a clinical and dysmorphologic evaluation of 194 individuals and review of the literature. Eur J Pediatr 172, 927–945.

3. Portnoï, M.-F. (2009). Microduplication 22q11.2: A new chromosomal syndrome. Eur J Med Genet 52, 88–93.

4. Verbesselt, J., Zink, I., Breckpot, J., and Swillen, A. (2022). Cross[sectional and longitudinal findings in patients with proximal 22q11.2 duplication: A retrospective chart study. Am J Med Genet A 188, 46–57.

5. Yobb, T.M., Somerville, M.J., Willatt, L., Firth, H. v., Harrison, K., MacKenzie, J., Gallo, N., Morrow, B.E., Shaffer, L.G., Babcock, M., et al. (2005). Microduplication and Triplication of 22q11.2: A Highly Variable Syndrome. The American Journal of Human Genetics 76, 865–876.

6. Savoia, A., Kunishima, S., de Rocco, D., Zieger, B., Rand, M.L., Pujol-Moix, N., Caliskan, U., Tokgoz, H., Pecci, A., Noris, P., et al. (2014). Spectrum of the Mutations in Bernard-Soulier Syndrome. Hum Mutat 35, 1033–1045.

7. Nunes, N., Zamariolli, M., Dantas, A.G., Cola, P., de Agostinho Júnior, F., Piazzon, F.B., Meloni, V.A., and Melaragno, M.I. (2022). CEDNIK syndrome in a Brazilian patient with compound heterozygous pathogenic variants. Eur J Med Genet 65, 104440.

8. Kingdom, R., and Wright, C.F. (2022). Incomplete Penetrance and Variable Expressivity: From Clinical Studies to Population Cohorts. Front Genet 13,.

9. Wright, C.F., West, B., Tuke, M., Jones, S.E., Patel, K., Laver, T.W., Beaumont, R.N., Tyrrell, J., Wood, A.R., Frayling, T.M., et al. (2019). Assessing the Pathogenicity, Penetrance, and Expressivity of Putative Disease-Causing Variants in a Population Setting. The American Journal of Human Genetics 104, 275–286.

10. Auwerx, C., Lepamets, M., Sadler, M.C., Patxot, M., Stojanov, M., Baud, D., Mägi, R., Porcu, E., Reymond, A., Kutalik, Z., et al. (2022). The individual and global impact of copy-number variants on complex human traits. The American Journal of Human Genetics 109, 647–668.

11. Davies, R.W., Fiksinski, A.M., Breetvelt, E.J., Williams, N.M., Hooper, S.R., Monfeuga, T., Bassett, A.S., Owen, M.J., Gur, R.E., Morrow, B.E., et al. (2020). Using common genetic variation to examine phenotypic expression and risk prediction in 22q11.2 deletion syndrome. Nat Med 26, 1912–1918.

12. Bycroft, C., Freeman, C., Petkova, D., Band, G., Elliott, L.T., Sharp, K., Motyer, A., Vukcevic, D., Delaneau, O., O’Connell, J., et al. (2018). The UK Biobank resource with deep phenotyping and genomic data. Nature 562, 203–209.

13. Köhler, S., Gargano, M., Matentzoglu, N., Carmody, L.C., Lewis-Smith, D., Vasilevsky, N.A., Danis, D., Balagura, G., Baynam, G., Brower, A.M., et al. (2021). The Human Phenotype Ontology in 2021. Nucleic Acids Res 49, D1207–D1217.

14. Wu, P., Gifford, A., Meng, X., Li, X., Campbell, H., Varley, T., Zhao, J., Carroll, R., Bastarache, L., Denny, J.C., et al. (2019). Mapping ICD-10 and ICD-10-CM Codes to Phecodes: Workflow Development and Initial Evaluation. JMIR Med Inform 7, e14325.

15. Bastarache, L., Hughey, J.J., Hebbring, S., Marlo, J., Zhao, W., Ho, W.T., van Driest, S.L., McGregor, T.L., Mosley, J.D., Wells, Q.S., et al. (2018). Phenotype risk scores identify patients with unrecognized Mendelian disease patterns. Science (1979) 359, 1233–1239.

16. Macé, A., Tuke, M.A., Beckmann, J.S., Lin, L., Jacquemont, S., Weedon, M.N., Reymond, A., and Kutalik, Z. (2016). New quality measure for SNP array based CNV detection. Bioinformatics 32, 3298–3305.

17. Gao, X., Starmer, J., and Martin, E.R. (2008). A multiple testing correction method for genetic association studies using correlated single nucleotide polymorphisms. Genet Epidemiol 32, 361–369.

18. Porcu, E., Rüeger, S., Lepik, K., Santoni, F.A., Reymond, A., and Kutalik, Z. (2019). Mendelian randomization integrating GWAS and eQTL data reveals genetic determinants of complex and clinical traits. Nat Commun 10, 3300.

19. Võsa, U., Claringbould, A., Westra, H.-J., Bonder, M.J., Deelen, P., Zeng, B., Kirsten, H., Saha, A., Kreuzhuber, R., Yazar, S., et al. (2021). Large-scale cis- and trans-eQTL analyses identify thousands of genetic loci and polygenic scores that regulate blood gene expression. Nat Genet 53, 1300–1310.

20. Crawford, K., Bracher-Smith, M., Owen, D., Kendall, K.M., Rees, E., Pardiñas, A.F., Einon, M., Escott-Price, V., Walters, J.T.R., O’Donovan, M.C., et al. (2019). Medical consequences of pathogenic CNVs in adults: analysis of the UK Biobank. J Med Genet 56, 131–138.

21. Owen, D., Bracher-Smith, M., Kendall, K.M., Rees, E., Einon, M., Escott-Price, V., Owen, M.J., O’Donovan, M.C., and Kirov, G. (2018). Effects of pathogenic CNVs on physical traits in participants of the UK Biobank. BMC Genomics 19, 867.

22. Aguirre, M., Rivas, M.A., and Priest, J. (2019). Phenome-wide Burden of Copy-Number Variation in the UK Biobank. The American Journal of Human Genetics 105, 373–383.

23. Kendall, K.M., Bracher-Smith, M., Fitzpatrick, H., Lynham, A., Rees, E., Escott-Price, V., Owen, M.J., O’Donovan, M.C., Walters, J.T.R., and Kirov, G. (2019). Cognitive performance and functional outcomes of carriers of pathogenic copy number variants: analysis of the UK Biobank. The British Journal of Psychiatry 214, 297–304.

24. Kendall, K.M., Rees, E., Bracher-Smith, M., Legge, S., Riglin, L., Zammit, S., O’Donovan, M.C., Owen, M.J., Jones, I., Kirov, G., et al. (2019). Association of Rare Copy Number Variants With Risk of Depression. JAMA Psychiatry 76, 818.

25. Voll, S.L., Boot, E., Butcher, N.J., Cooper, S., Heung, T., Chow, E.W.C., Silversides, C.K., and Bassett, A.S. (2017). Obesity in adults with 22q11.2 deletion syndrome. Genetics in Medicine 19, 204–208.

26. Loid, P., Pekkinen, M., Mustila, T., Tossavainen, P., Viljakainen, H., Lindstrand, A., and Mäkitie, O. (2022). Targeted Exome Sequencing of Genes Involved in Rare CNVs in Early-Onset Severe Obesity. Front Genet 13,.

27. Campbell, I.M., Crowley, T.B., Jobaliya, C., Bailey, A., McGinn, D.E., Gaiser, K., Bassett, A., Gur, R.E., Morrow, B., Emanuel, B.S., et al. (2022). Platelet findings in 22q11.2 Deletion Syndrome correlate with disease manifestations but do not correlate with GP1b surface expression. MedRxiv 2022.06.10.22276258.

28. Hierck, B.P., Molin, D.G.M., Boot, M.J., Poelmann, R.E., and Gittenberger-De Groot, A.C. (2004). A Chicken Model for DGCR6 as a Modifier Gene in the DiGeorge Critical Region. Pediatr Res 56, 440–448.

29. Yu, A., Turbiville, D., Xu, F., Ray, J.W., Britt, A.D., Lupo, P.J., Jain, S.K., Shattuck, K.E., Robinson, S.S., and Dong, J. (2019). Genotypic and phenotypic variability of 22q11.2 microduplications: An institutional experience. Am J Med Genet A 179, 2178–2189.

30. Courtens, W., Schramme, I., and Laridon, A. (2008). Microduplication 22q11.2: A benign polymorphism or a syndrome with a very large clinical variability and reduced penetrance?—Report of two families. Am J Med Genet A 146A, 758–763.

31. Verbesselt, J., Zink, I., Breckpot, J., and Swillen, A. (2022). Cross[sectional and longitudinal findings in patients with proximal 22q11.2 duplication: A retrospective chart study. Am J Med Genet A 188, 46–57.

32. Fry, A., Littlejohns, T.J., Sudlow, C., Doherty, N., Adamska, L., Sprosen, T., Collins, R., and Allen, N.E. (2017). Comparison of Sociodemographic and Health-Related Characteristics of UK Biobank Participants With Those of the General Population. Am J Epidemiol 186, 1026–1034.

33. Kirov, G., Rees, E., Walters, J.T.R., Escott-Price, V., Georgieva, L., Richards, A.L., Chambert, K.D., Davies, G., Legge, S.E., Moran, J.L., et al. (2014). The Penetrance of Copy Number Variations for Schizophrenia and Developmental Delay. Biol Psychiatry 75, 378–385.

34. Olsen, L., Sparsø, T., Weinsheimer, S.M., dos Santos, M.B.Q., Mazin, W., Rosengren, A., Sanchez, X.C., Hoeffding, L.K., Schmock, H., Baekvad-Hansen, M., et al. (2018). Prevalence of rearrangements in the 22q11.2 region and population-based risk of neuropsychiatric and developmental disorders in a Danish population: a case-cohort study. Lancet Psychiatry 5, 573–580.

35. Kingdom, R., Tuke, M., Wood, A., Beaumont, R.N., Frayling, T.M., Weedon, M.N., and Wright, C.F. (2022). Rare genetic variants in genes and loci linked to dominant monogenic developmental disorders cause milder related phenotypes in the general population. The American Journal of Human Genetics 109, 1308–1316.

