## Supplementary Figure for "The impact of 22q11.2 copy number variants on human traits in the general population"

**Supplemental Figures and legends**

**Supplementary Figure 1 | 22q11.2 CNVs and significant continuous traits. Top:** The negative logarithm of the association p-value for the CNV-trait association for the model indicated in the legend is plotted against the 22q11.2 genomic region. Each point represents a CNV proxy probe. The red dashed line indicates significance threshold ($p < 6.5 \times 10^{-5}$). **Bottom:** Gray bars represent low copy-repeat region (LCR) A-D, as well as the 90 genes contained in the region. The 24 genes used for trait selection are labeled in black.

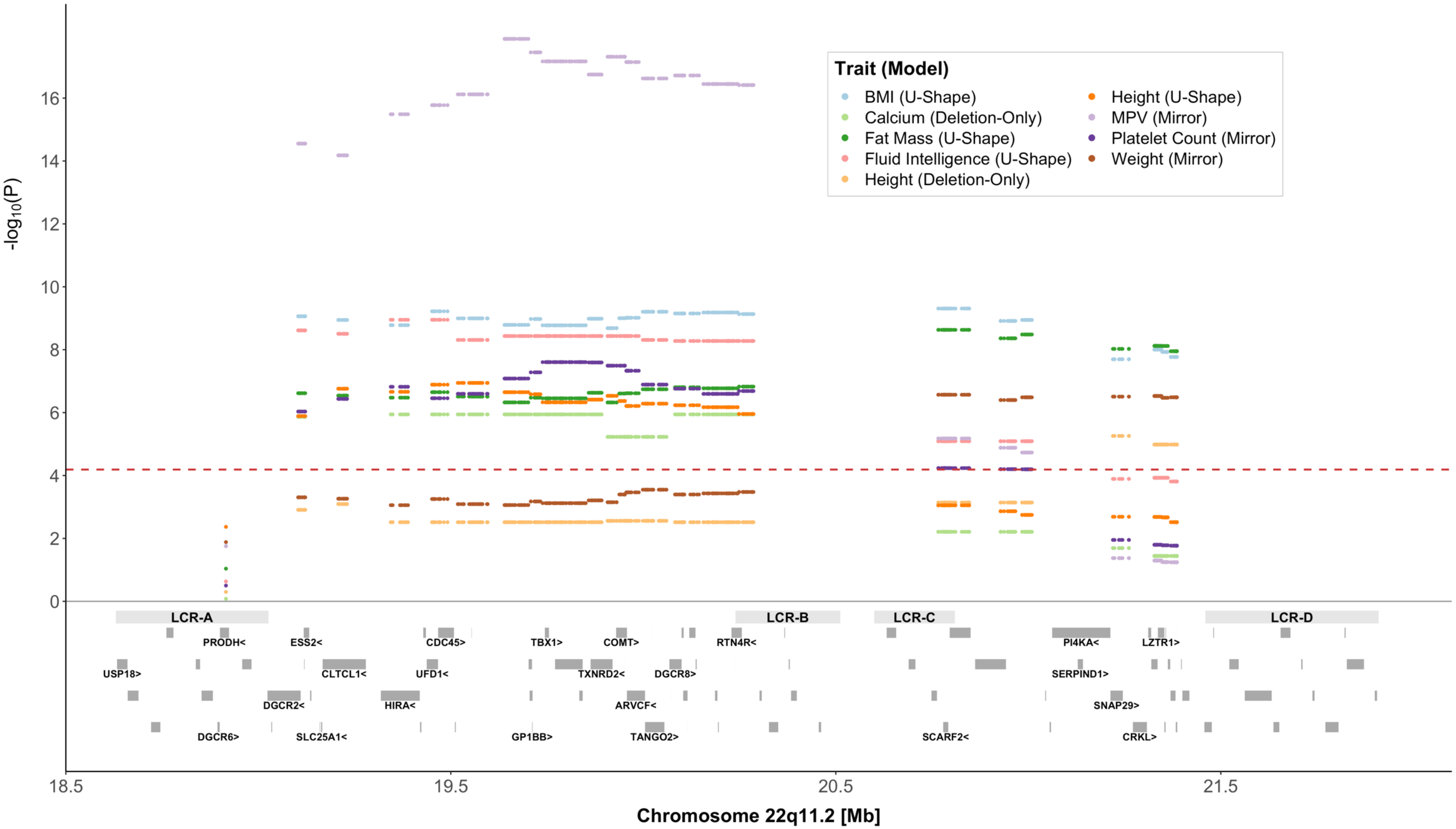
Trait (Model)
BMI (U-Shape)
Calcium (Deletion-Only)
Fat Mass (U-Shape)
Fluid Intelligence (U-Shape)
Height (Deletion-Only)
Height (U-Shape)
MPV (Mirror)
Platelet Count (Mirror)
Weight (Mirror)
-log10(P)
Chromosome 22q11.2 [Mb]
18.5
19.5
20.5
21.5
LCR-A
LCR-B
LCR-C
LCR-D
PRODH<
ESS2<
CDC45>
TBX1>
COMT>
RTN4R<
PI4KA<
LZTR1>
USP18>
CLTCL1<
UFD1<
TXNRD2<
DGCR8>
SERPIND1>
DGCR2<
HIRA<
ARVCF<
SNAP29>
DGCR6>
SLC25A1<
GP1BB>
TANGO2>
SCARF2<
CRKL>

**Supplementary Figure 2 | 22q11.2 CNVs and significant binary traits. Top:** The negative logarithm of the association p-value for the CNV-trait association for the model indicated in the legend is plotted against the 22q11.2 genomic region. Each point represents a CNV proxy probe. The red dashed line indicates significance threshold ($p < 6.5 \times 10^{-5}$). **Bottom:** Gray bars represent low copy-repeat region (LCR) A-D, as well as the 90 genes contained in the region. The 24 genes linked to traits according to HPO are labeled in black.

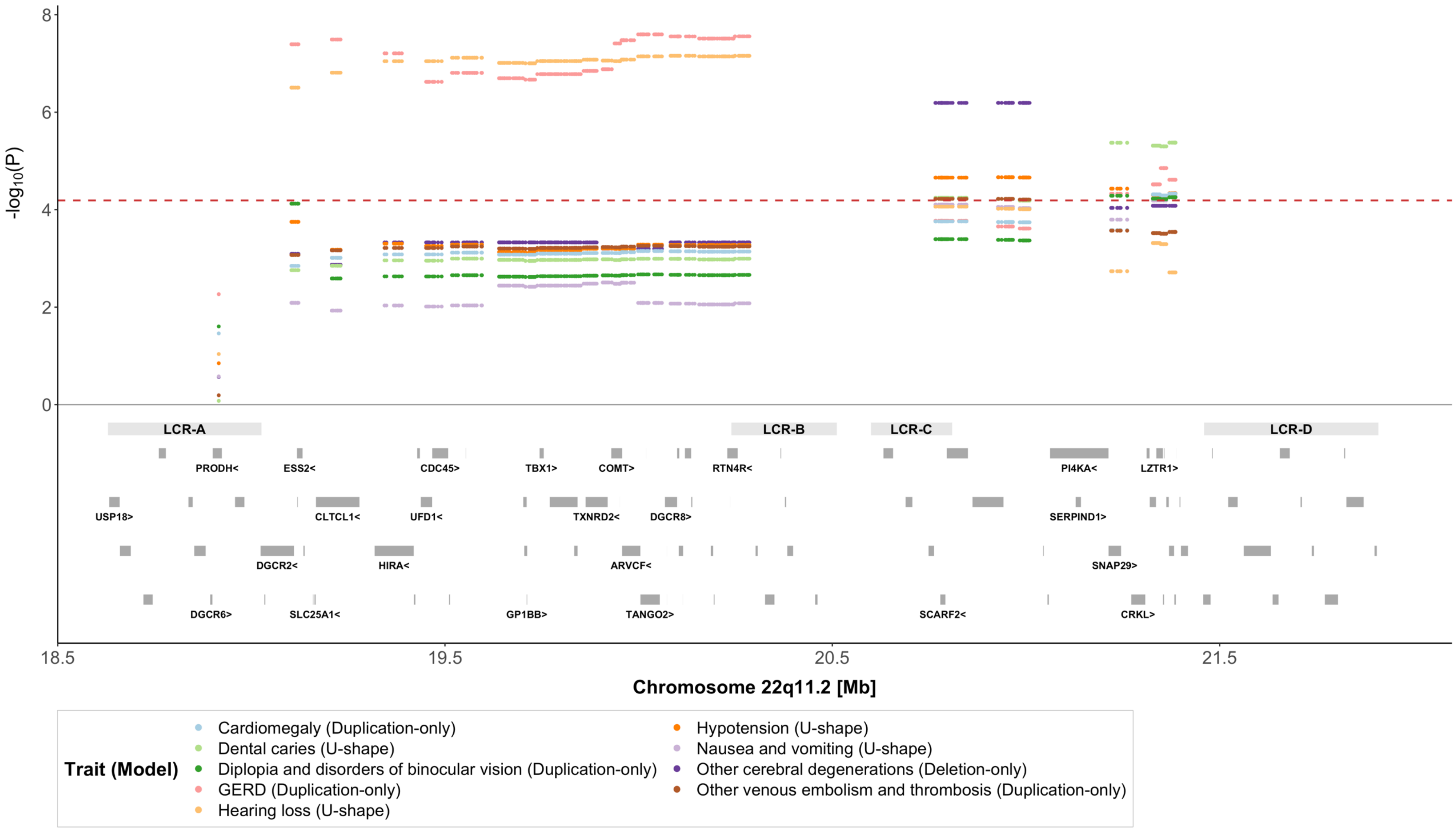

-log10(P)
8
6
4
2
0
LCR-A
LCR-B
LCR-C
LCR-D
PRODH<
ESS2<
CDC45>
TBX1>
COMT>
RTN4R<
PI4KA<
LZTR1>
USP18>
CLTCL1<
UFD1<
TXNRD2<
DGCR8>
SERPIND1>
DGCR2<
HIRA<
ARVCF<
SNAP29>
DGCR6>
SLC25A1<
GP1BB>
TANGO2>
SCARF2<
CRKL>
18.5
19.5
20.5
21.5
Chromosome 22q11.2 [Mb]
Trait (Model)
Cardiomegaly (Duplication-only)
Dental caries (U-shape)
Diplopia and disorders of binocular vision (Duplication-only)
GERD (Duplication-only)
Hearing loss (U-shape)
Hypotension (U-shape)
Nausea and vomiting (U-shape)
Other cerebral degenerations (Deletion-only)
Other venous embolism and thrombosis (Duplication-only)

**Supplementary Figure 3 | HPO terms linked to 22q11.2 genes through different genetic variants.** Barplot showing the number of HPO terms used in this study linked to conditions caused by 22q11.2 CNVs (green), genetic variants affecting a single gene in the region (pink) or linked to the 22q11.2 gene loci (orange).

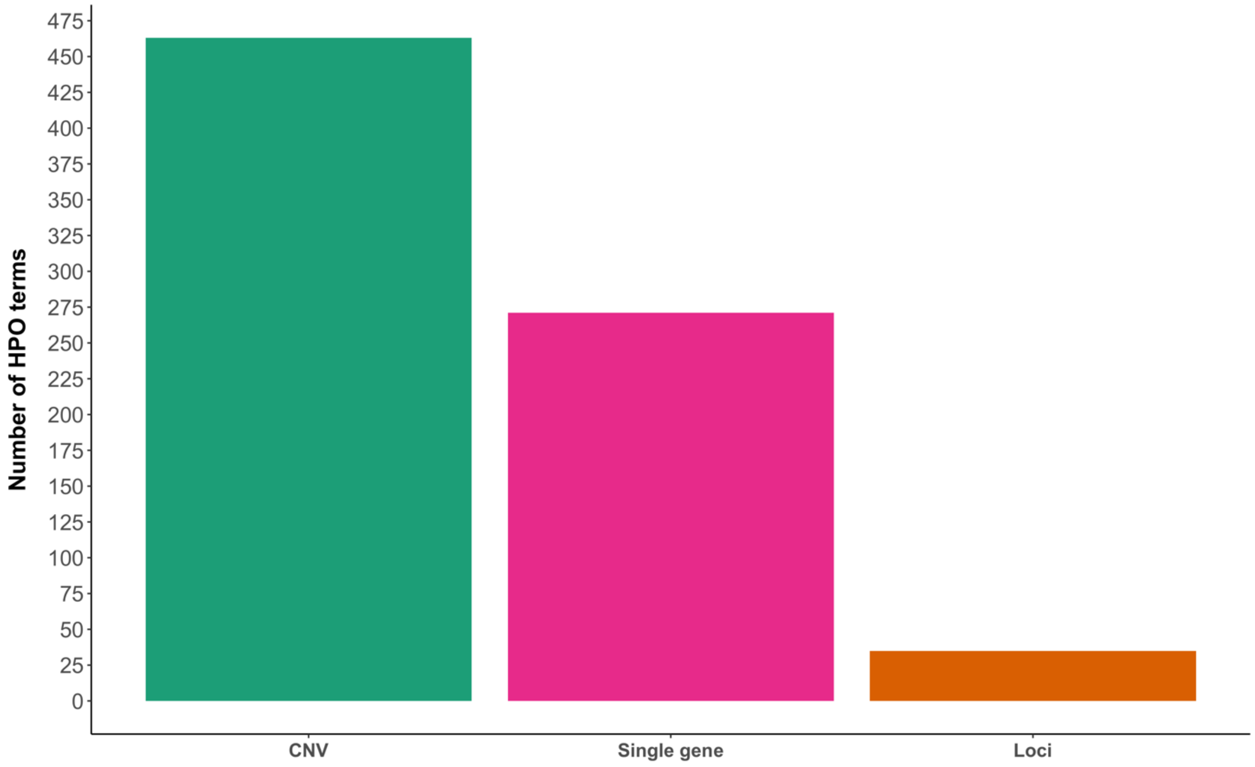
